# Performance evaluation of nine reference centers for effective surveillance of *Leishmania*-infected Phlebotomine sand flies and basis for technical recommendations

**DOI:** 10.1101/2024.09.19.24313901

**Authors:** Jorian Prudhomme, Aymeric Delabarre, Bulent Alten, Umut Berberoglu, Eduardo Berriatua, Gioia Bongiorno, José Manuel Cristovao, Maya Davidovich-Cohen, Trentina Di Muccio, Ozge Erisoz Kasap, Eleonora Fiorentino, Oscar D. Kirstein, Edwin Kniha, Carla Maia, Mesut Mungan, Clara Muñoz-Hernández, Muhammed Nalçaci, Gizem Oguz Kaskan, Yusuf Ozbel, Seray Ozensoy Toz, Ricardo Parreira, Katharina Platzgummer, Ceylan Polat, José Risueño, Liora Studentsky, Gamze Varol, Julia Walochnik, Kardelen Yetişmiş, Florence Robert-Gangneux

## Abstract

**Background:** Leishmaniasis, caused by *Leishmania* protozoan parasites transmitted by Phlebotomine sand flies, is a significant public health concern in the Mediterranean basin. Effective monitoring of *Leishmania*-infected sand flies requires standardized tools for comparing their distribution and infection prevalence. Consistent quantitative PCR (qPCR) conditions and efficient DNA extraction protocols are crucial for reliable results over time and across regions. However, there is currently a lack of technical recommendations for *Leishmania* DNA detection, which needs to be addressed. This study aimed to compare various DNA extraction protocols and conduct a qPCR based External Quality Assessment (EQA) through a multicenter study involving nine reference laboratories.

**Methodology/Principal findings:** EQA samples were prepared using *Leishmania infantum* and *L. major* strains, at different concentration from 10^1^ to 10^4^ parasites/mL and distributed to participating centers. All centers, except one, detected all *Leishmania* concentrations, demonstrating diagnostic proficiency. The ability to detect low concentrations highlighted the robustness of the qPCR assay used, although Cq value variations suggested differences in sensitivity due to technical capabilities and/or extraction kit performances.

Reported comparative analysis of seven DNA extraction methods identified the EZ1 DSP Virus® Kit and QIAamp® DNA mini-kit as the most efficient, supporting their use for standardized protocols. The study also evaluated the impact of lyophilization and shipment conditions, finding no compromise in *Leishmania* detection, despite slight Cq value variations. In addition to EQA samples, experimentally infected sand fly have been included to mimic sample field condition. All centers detected positive samples, with variable Cq values, reflecting differences in individual infection load.

**Conclusion and significance:** Overall, the study underscores the importance of standardized protocols and continuous quality assurance to maintain high diagnostic validity, crucial for effective surveillance of leishmaniasis, especially in field settings with low infection densities. Continuous training and calibration are essential to ensure uniform diagnostic performance across laboratories, enhancing epidemiological surveillance and disease control strategies.

**Author Summary:** Leishmaniasis is a disease caused by *Leishmania* parasites, transmitted by sand flies, and poses a major health risk in the Mediterranean region. Monitoring the spread of nfected sand flies is crucial for controlling the disease. This study focused on improving the methods used to detect *Leishmania* in sand flies by comparing different DNA extraction techniques and assessing the accuracy of these methods across nine reference laboratories. All centers, except one, efficiently detected all *Leishmania* concentrations, demonstrating proficiency in diagnostic protocols. Moreover, we found that two specific DNA extraction kits, the EZ1 DSP Virus® Kit and QIAamp® DNA mini-kit, were the most effective for *Leishmania* detection. We also tested how sample preparation and shipping conditions affected the results, ensuring that our methods would work in real-world settings. Even under these conditions, the detection methods proved reliable. This work helps to standardize the detection of *Leishmania*, making surveillance more accurate and consistent. Continuous training and calibration are essential to ensure uniform diagnostic performance across laboratories, enhancing epidemiological surveillance and disease control strategies

## Introduction

Leishmaniasis is a disease caused by parasites of the genus *Leishmania* and transmitted by the bite of Phlebotomine sand flies. This parasitic infection is endemic in territories around the Mediterranean basin, where it represents a significant public health concern (1). Leishmaniasis clinical manifestations are diverse, ranging from cutaneous lesions, which may cause disfiguring ulcers on exposed parts of the body, to visceral disease with infiltration of the lymph nodes, spleen, liver, and bone marrow, causing pancytopenia and being fatal if untreated (2, 3). Despite the significant health impact of leishmaniasis worldwide, there is no substantial evidence indicating a rise in the incidence of autochthonous human cases in Europe. However, the disease remains often underreported, leading to a possible underestimation of its true burden (4). Hypothesis confirmed by a notable increase of autochthonous canine leishmaniasis cases (5). Furthermore, the geographical distribution of leishmaniasis is changing. New foci of infection are emerging in areas previously considered non-endemic, while old foci are re-emerging (5–7). Triggering factors contributing to these evolving scenario include climate change, which affects the distribution and behavior of sand fly vectors, increased movement of people and animals and trade activities, which can cause parasite introduction into new areas. Overall, the dynamic epidemiology of leishmaniasis in Europe underscores the need for vigilant surveillance and reporting systems.

Effective epidemiologic surveillance of *Leishmania*-infected sand flies could be considered an essential tool for understanding and controlling the spread of leishmaniasis. To achieve this goal, standardized protocols are required to accurately compare the distribution areas and the prevalence of sand fly infection. Real-time quantitative PCR (qPCR), which is a sensitive and specific method for detecting *Leishmania* DNA, plays a crucial role in this process (8). Utilizing consistent amplification conditions and similar extraction protocols across different laboratories is vital for ensuring that the results are comparable over time and across various geographical areas (9). Moreover, reliable data merged from different regions can help in mapping the spread of the disease and in understanding the factors driving its transmission, such as climate change, urbanization, and movements of infected hosts and vectors (10).

Despite the importance of these techniques, there is currently a lack of evaluation in nucleic acid extraction and qPCR techniques for *Leishmania* diagnosis. This deficiency represents a significant gap in the epidemiologic surveillance framework. Without standardized and validated methods, the reliability of data collected from different studies can be compromised, making it difficult to draw accurate comparisons and conclusions. Laboratory efficiency has been compared throughout European countries for the diagnosis of other parasitic or fungal diseases, such as toxoplasmosis (11), histoplasmosis (9) and *Pneumocystis* pneumonia (12). Regarding leishmaniasis, a European study (13) has previously compared the accuracy of species identification by molecular methods, but no such initiative has been implemented for evaluating *Leishmania* detection by qPCR.

In this context, the objectives of this study were twofold: (i) to analyze the performance of various DNA extraction protocols for detecting *L. infantum* and *L. major*, by qPCR and (ii) to conduct an External Quality Assessment (EQA) through a multicenter study involving nine reference laboratories, participating in the European project CLIMOS (http://www.climos-project.eu) which collects data on sand fly infections. This study aimed at ensuring the reliability and comparability of *Leishmania* detection methods from sand flies across different regions and laboratories, thereby enhancing the accuracy of epidemiological surveillance and contributing to more effective disease control strategies.

## Methods

### Participants and study design

The Laboratory of Parasitology of Rennes University/Institut National de la Santé et de la Recherche Médicale (INSERM) (Rennes, France), which is a reference laboratory for the diagnosis of leishmaniasis and other parasitic and fungal infections, was in charge of developing standard operation procedures (SOP) for *Leishmania* extraction from sand flies and oversaw the implementation of the EQA program for CLIMOS. The Rennes Lab prepared the EQA samples and evaluated the various extraction methods used by eight European and non-European laboratories involved in the project, located in 6 countries, including: the so-called reference center (INSERM, Rennes, France), Ege University (EGE, Izmir, Turkey), Hacettepe Universitesi (HACETTEPE, Ankara, Turkey), Institute of Hygiene and Tropical Medicine, Universidade Nova de Lisboa (UNL, Lisboa, Portugal), Jerusalem Public Health laboratories, Ministry of Health (IMOH, Jerusalem, Israel), Istituto Superiore di Sanita (ISS, Roma, Italy), Medizinische Universitaet Wien (MEDUNI VIENNA, Wien, Austria), Turkiye Cumhuriyeti Saglik Bakanligi (MOH, Ankara, Turkey) and Universidad de Murcia (UM, Murcia, Spain). The participating centers other than the reference center were designated as “Center 1 to Center 8”. The EQA program for *Leishmania* DNA extraction and qPCR analysis involved testing cultured parasites and experimentally infected sand flies.

### Sand fly samples

For all experiments and EQA samples, we used *Phlebotomus perniciosus* from well-adapted laboratory colonies. Sand flies were provided by Istituto Superiore di Sanita (ISS) (Roma, Italy) and Hacettepe Universitesi (HU) (Ankara, Turkey) for uninfected specimens, and by Charles University (CUNI) (Prague, Czech Republic) for experimentally infected ones (14).

Leishmania *strains and preparation of EQA samples*

Two strains of *Leishmania* were used, *L. infantum* #REN-12-02 and *L. major* #REN-22-02 (both cryopreserved at the Biological Resource Center of the Rennes University Hospital and Leishmaniasis Reference Center of Montpellier University Hospital) for the comparison of DNA extraction techniques, preparation of EQA samples and EQA validation. Promastigotes were maintained in an incubator at 26°C by weekly transfers in T25 flask containing M199 medium (Sigma) supplemented with 10% inactivated fetal calf serum, 1% HEPES, 1% Penicillin-Streptomycin, 1% hypoxanthin, 0.2% hemin, 0.1% biotin and 0.4% biopterin.

Five serial 1:10 dilutions of each strain containing 10^5^, 10⁴, 10³, 10² and 10¹ parasite/mL were prepared, starting with, 1 mL of homogenized broth culture. Dilutions were carried out in a 5% formalinized Dulbecco’s phosphate-buffered saline (DPBS) solution. Promastigotes were counted using the standardized KOVA cell chamber system, according to the protocol established by the supplier (Kova International, California, USA). To ensure accuracy, the counting was realized in 3 cells and by two different operators. Ready to used, parasite suspensions were aliquoted into 1.5 mL tubes and directly stored at -20 °C or lyophilized and stored at -20°C until use. A set of lyophilized samples was kept at room temperature for 3 weeks to evaluate the impact of storage conditions on qPCR results.

### EQA sample processing

All centers received a panel of ten EQA samples, consisting of eight lyophilized (*i.e. L. major* and *L. infantum* at 10⁴, 10³, 10² and 10¹ parasite/mL) and two liquid samples (*i.e.* one uninfected and one experimentally infected sand fly in 200 µL of PBS). At reception, samples were stored at −20°C until further testing. Lyophilized samples were rehydrated with 200 µL of PCR-quality water and sand fly samples were processed like any sand fly collected from the field for analysis, *i.e.* grinded in a final volume of 700 µL of PBS and incubated at 56°C during 2 hours with proteinase K. Then, extractions were realized with an extraction volume of 400 µL (200 µL of EQA and 200 µL lysis buffer) and an elution volume between 50 and 90 µL according to each center technique and equipment (Table 1).

**Table 1.** Methods used for Leishmania nucleic acids extraction and detection in the different center.

| Center | Extraction kit | Extraction device | Elution volume | Amplification device |
| --- | --- | --- | --- | --- |
| Reference center | EZ1 DSP Virus® Kit | EZ1 extraction device | 90 | StepOne Real-time PCR System |
| 1 | RSC Blood DNA® | Maxwell RSC® 16 instrument | 50 | Biorad iQ5 |
| 2 | Allprep® DNA/RNA micro kit | Handmade | 90 | Biorad CFX96 Real-time system |
| 3 | QIAmp® viral RNA mini kit | Handmade | 90 | Rotor Gene 3000 |
| 4 | Kit 1: RSC Viral TNA® | Maxwell RSC® 16 instrument | 90 | QuantStudio™ 5 Real-Time PCR |
|  | Kit 2: RSC Blood DNA® |  | 50 |  |
| 5 | Allprep® DNA/RNA mini kit | Handmade | 90 | StepOnePlus Real-time PCR System |
| 6 | EZ1&2 virus mini® kit | EZ1 extraction device | 90 | Biorad CFX96 Real-time system |
| 7 | DNeasy® Blood & Tissue Kit | Handmade | 90 | Rotor-Gene Q |
| 8 | Mag-Bind® Blood & Tissue DNA Kit | TissueLyser II | 90 | Biorad CFX96 Real-time system |

All partners employed the same qPCR method (15) based on the amplification of a kinetoplast DNA (kDNA) minicircle sequence with primers and Taqman probe: 5’-CTT-TTC-TGG-TCC-TCC-GGG-TAGG, 5’-CCA-CCC-GGC-CCT-ATT-TTA-CAC-CAA and 5’ FAM-TTT-TCG-CAG-AAC-GCC-CCT-ACC-CGC-3’ TAMRA, respectively, provided by the reference center. Each 25 μL qPCR reaction mix included 5 μL of DNA sample, 12.5 μL of TaqMan Universal Master Mix 2X and a final concentration of 0.5 μM of primers and 0.2 μM of probe. DNA was amplified using the following conditions: initial step at 95 °C for 10 min, followed by 45 cycles of 15 sec at 95°C and 1 min at 60°C. Participating centers used their own qPCR device (Table 1), realized the amplification in triplicates and included their own positive and negative controls. The qPCR threshold cycle (Cq) defined as the cycle at which near logarithmic product amplification takes place, was used as a semi-quantitative measure of parasite DNA concentration (16).

### Comparison of DNA extraction techniques

As the amplification method was the same for all participating centers, we suspected that variations might appear related, at least partly, to the extraction method used. Therefore, we undertook the evaluation of seven extraction methods, including some used by the participating centers (Table 1), and additional ones which were designed to purify total nucleic acids and could offer the opportunity to detect simultaneously *Phlebovirus*, also transmitted by sand flies.

Seven kits were compared for *Leishmania* DNA extraction, including the following five manual extraction kits: EZ1 DSP Virus® Kit using EZ1 extraction device (Qiagen, Hilden, Germany), RNeasy® mini kit (Qiagen, Hilden, Germany), QIAamp® DNA mini kit (Qiagen, Hilden, Germany), Allprep® DNA/RNA mini kit (Qiagen, Hilden, Germany), QIAamp® viral RNA mini kit (Qiagen, Hilden, Germany) and two automated extraction kits: RSC Viral TNA® (Promega, Southampton, England) and RSC Blood DNA® (Promega, Southampton, England) using Maxwell RSC® 48 instrument (Promega). Amplifications were realized using a QuantStudio™ 5 Real-Time PCR System (Applied Biosystems®, Thermo Fisher Scientific®, Villebon-sur-Yvette, France). Liquid samples, containing parasite suspensions, aliquoted in small vials and stored at -20°C were used for this evaluation, to avoid possible variations due to the lyophilization process and reconstitution. Extractions were performed in triplicate from 3 independent vials of each concentration, according to the manufacturer’s instructions. Amplifications were also performed in triplicates, using 5 µL of DNA in a final volume of 25 µL as described above.

### Assessment of external conditions potentially influencing individual performances

First, the variability of *Leishmania* spp. DNA detection of infected sand flies using seven individuals extracted with the same kit was assessed (EZ1 DSP Virus ® Kit using EZ1). Then, the process (*i.e.* lyophilization and shipment conditions) was tested through three experiments. The impact of lyophilization was assessed by DNA extraction of *L. infantum* and *L. major* aliquots at four concentrations (10⁴, 10³, 10² and 10¹), before and after lyophilization. Second, to ensure there was no impact of shipment conditions on sample quality, results obtained with samples stored at room temperature (RT) for 3 weeks and samples stored at –20°C for the same time, before DNA extraction and amplification were compared. Third, the potential inhibitory effect of sand fly DNA on the detection of low amounts of *Leishmania* DNA was tested. For this purpose, pools of sand flies (30 individuals, 15 males and 15 females) were spiked with 100 or 1000 *Leishmania* (*L. major* or *L. infantum*) promastigotes and grinded in a final volume of 700 µL of PBS, mimicking usual practice for field studies. The same numbers of *Leishmania* without sand flies were used as controls. Homogenates were submitted to a 2-hour heating step with proteinase K at 56°C before DNA extraction. Two hundred µL were used for DNA extraction using EZ1 DSP Virus® Kit (Qiagen, Hilden, Germany) according to the manufacturer’s instructions and eluted in 90 µL of elution buffer.

Amplification was carried out using a QuantStudio™ 5 Real-Time PCR device (Applied Biosystems®). Extractions and amplifications were performed in triplicates.

### Statistical analysis

Results were presented as mean ±SD of quantification cycle (Cq) values of amplification of each parasite concentration for each center. They were compared using two-way ANOVA or mixed-effects analyses (if missing data were present) and a Tukey’s multiple comparisons test as post-hoc analyses. All analyses and graphics were realized with GraphPad ® Prism Software version 9.

## Results

### Multicenter qPCR analysis of EQA samples

Results of DNA amplification by qPCR of the two *Leishmania* species at four different concentrations, obtained by the eight centers, are provided in Fig 1. All centers correctly amplified positive samples except center 6 which failed to amplify *L. infantum* at 10^1^/mL concentration. For *L. infantum*, mean Cq values across centers ranged from approximately 26 to 32, 29 to 32 and 31 to 36 for the 10⁴/mL, 10³/mL and 10²/mL parasite concentrations, respectively (Fig 1A). Similarly, the mean Cq values obtained for *L. major* showed notable inter-center variations, with overall higher mean Cq values compared to those obtained in the *L. infantum* assay (Fig 1B). The mean Cq values ranged from 26 to 38 for samples with 10⁴/mL parasites and from approximately 30 to 45 for samples with 10²/mL parasites. At the lowest *L. major* concentration of 10¹/mL parasites, Cq values ranged from 33 to 43. All participating centers also accurately detected the samples containing a *L. infantum* experimentally infected sand fly, with variable mean Cq ranging from 22 to 42 (Fig 2).

**Fig 1.**
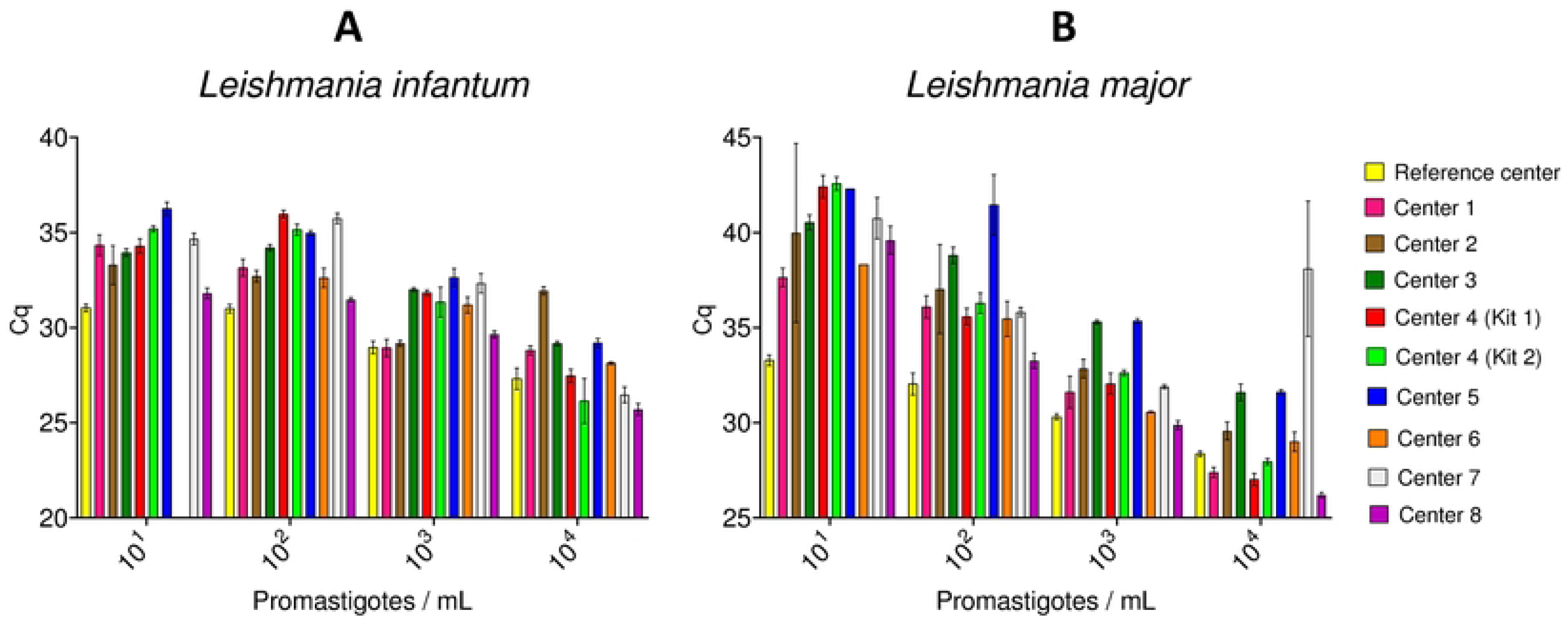
PCR results for EQA samples of *Leishmania infantum* (A) and *Leishmania major* (B) promastigotes at indicated concentrations by participating centers (mean Cq ± SD of triplicate amplification).

**Fig 2.**
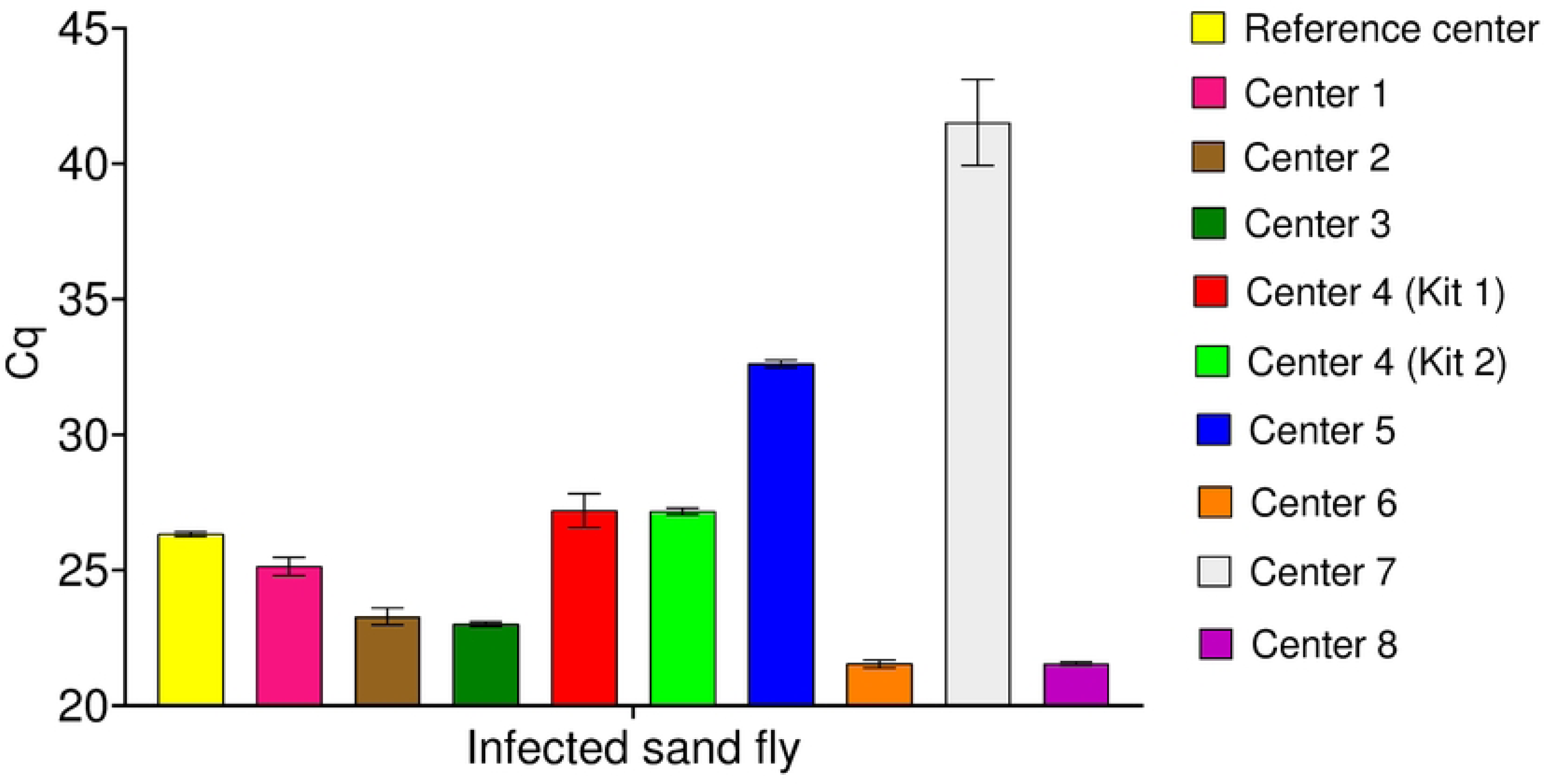
Detection of *Leishmania infantum* DNA from an infected sand fly. Real-time quantitative PCR results of participating centers (mean Cq ± SD of triplicate amplification).

### Comparison of extraction protocols

For both *L. infantum* and *L. major*, the use of EZ1 DSP Virus® Kit, QIAamp® DNA mini kit, Allprep® DNA/RNA mini kit, and RSC Blood® DNA yielded the lowest Cq values for all concentrations, indicating they are the most efficient kits in extracting DNA from *Leishmania*. Even though these four kits were associated with low Cq values, extraction with EZ1 DSP Virus Kit offered the best overall efficiency since mean Cq values were significantly lower than those for other kits at most *L. infantum* and *L. major* concentrations (Tables 2 & 3). Conversely, RNeasy® mini kit, QIAamp viral RNA® mini kit, and RSC Viral TNA® showed higher Cq values, indicating lower efficiency in extracting DNA. The detailed comparisons between the various extraction kits for *L. infantum* are depicted in Fig 3A and p-values are summarized in Table 2, and those for *L. major* are illustrated in Fig 3B and summarized in Table 3.

**Fig 3.**
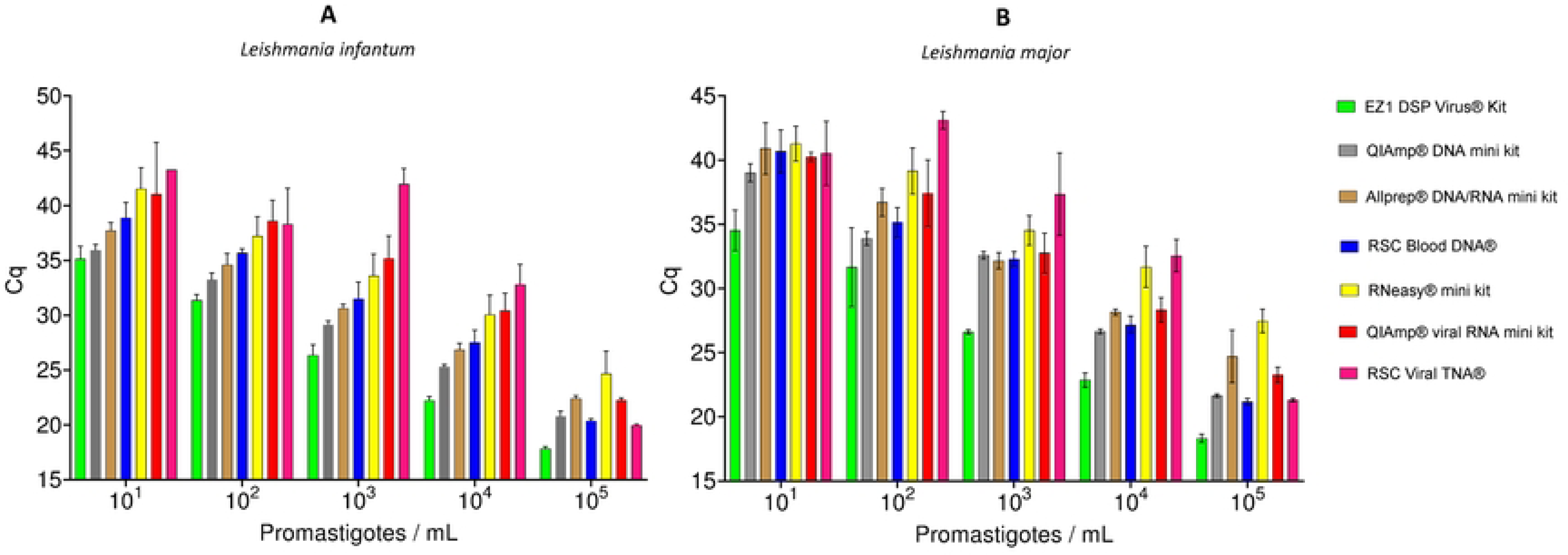
Efficiency of *Leishmania infantum* (A) and *Leishmania major* (B) detection following nucleic acids extraction using various kits. Results show mean Cq ± SD of triplicate amplification for each parasite concentration.

**Table 2.** *Detailed P-value for Tukey’s multiple comparisons test between extraction kits for* Leishmania infantum.

| Kit compared \ Concentration | Concentration |  |  |  |  |
| --- | --- | --- | --- | --- | --- |
|  | 10<sup>1</sup> | 10<sup>2</sup> | 10<sup>3</sup> | 10<sup>4</sup> | 10<sup>5</sup> |
| EZ1 DSP Virus® Kit vs RSC Blood DNA® | 0.0003 | <0.0001 | <0.0001 | <0.0001 | <0.0001 |
| EZ1 DSP Virus® Kit vs QIAamp® viral RNA mini kit | 0.5302 | <0.0001 | <0.0001 | <0.0001 | <0.0001 |
| EZ1 DSP Virus® Kit vs RNeasy® mini kit | 0.0014 | <0.0001 | <0.0001 | <0.0001 | 0.0001 |
| EZ1 DSP Virus® Kit vs RSC Viral TNA® | NA | 0.0275 | <0.0001 | <0.0001 | <0.0001 |
| EZ1 DSP Virus® Kit vs Allprep® DNA/RNA mini kit | 0.0009 | <0.0001 | <0.0001 | <0.0001 | <0.0001 |
| EZ1 DSP Virus® Kit vs QIAamp® DNA mini kit | 0.5452 | <0.0001 | 0.0001 | <0.0001 | <0.0001 |
| RSC Blood DNA® vs QIAamp® viral RNA mini kit | 0.9680 | 0.0188 | 0.0086 | 0.0067 | <0.0001 |
| RSC Blood DNA® vs RNeasy® mini kit | 0.1585 | 0.2685 | 0.2256 | 0.0366 | 0.0028 |
| RSC Blood DNA® vs RSC Viral TNA® | NA | 0.5402 | <0.0001 | <0.0001 | 0.0048 |
| RSC Blood DNA® vs Allprep® DNA/RNA mini kit | 0.3833 | 0.1166 | 0.6613 | 0.7066 | <0.0001 |
| RSC Blood DNA® vs QIAamp® DNA mini kit | 0.0019 | <0.0001 | 0.0167 | 0.0030 | 0.2812 |
| QIAamp® viral RNA mini kit vs RNeasy® mini kit | >0.9999 | 0.6866 | 0.6467 | 0.9988 | 0.0754 |
| QIAamp® viral RNA mini kit vs RSC Viral TNA® | NA | >0.9999 | <0.0001 | 0.1162 | <0.0001 |
| QIAamp® viral RNA mini kit vs Allprep® DNA/RNA mini kit | 0.8537 | 0.0016 | 0.0016 | 0.0011 | 0.8189 |
| QIAamp® viral RNA mini kit vs QIAamp® DNA mini kit | 0.6189 | 0.0002 | 0.0002 | <0.0001 | <0.0001 |
| RNeasy® mini kit vs RSC Viral TNA® | NA | 0.9847 | <0.0001 | 0.0671 | 0.0016 |
| RNeasy® mini kit vs Allprep® DNA/RNA mini kit | 0.0317 | 0.0273 | 0.0214 | 0.0064 | 0.1029 |
| RNeasy® mini kit vs QIAamp® DNA mini kit | 0.0054 | 0.0012 | 0.0015 | 0.0004 | 0.0049 |
| RSC Viral TNA® vs Allprep® DNA/RNA mini kit | NA | 0.2586 | <0.0001 | <0.0001 | <0.0001 |
| RSC Viral TNA® vs QIAamp® DNA mini kit | NA | 0.0941 | <0.0001 | <0.0001 | 0.0106 |
| Allprep® DNA/RNA mini kit vs QIAamp® DNA mini kit | 0.0010 | 0.0525 | <0.0001 | 0.0002 | <0.0001 |

NA: Not available due to an excessive amount of missing data for some kits at low concentrations.

**Table 3.** *Detailed P-value for Tukey’s multiple comparisons test between extraction kits for* Leishmania major.

| Concentration<br>Kit compared | 10<sup>1</sup> | 10<sup>2</sup> | 10<sup>3</sup> | 10<sup>4</sup> | 10<sup>5</sup> |
| --- | --- | --- | --- | --- | --- |
| EZ1 DSP Virus® Kit vs RSC Blood DNA® | <0.0001 | 0.0924 | <0.0001 | <0.0001 | <0.0001 |
| EZ1 DSP Virus® Kit vs QIAamp® viral RNA mini kit | <0.0001 | 0.0084 | <0.0001 | <0.0001 | <0.0001 |
| EZ1 DSP Virus® Kit vs RNeasy® mini kit | <0.0001 | 0.0004 | <0.0001 | <0.0001 | <0.0001 |
| EZ1 DSP Virus® Kit vs RSC Viral TNA® | 0.4009 | <0.0001 | <0.0001 | <0.0001 | <0.0001 |
| EZ1 DSP Virus® Kit vs Allprep® DNA/RNA mini kit | <0.0001 | 0.0111 | <0.0001 | <0.0001 | 0.0001 |
| EZ1 DSP Virus® Kit vs QIAamp® DNA mini kit | 0.0001 | 0.4043 | <0.0001 | <0.0001 | <0.0001 |
| RSC Blood DNA® vs QIAamp® viral RNA mini kit | 0.9878 | 0.2849 | 0.9720 | 0.1075 | <0.0001 |
| RSC Blood DNA® vs RNeasy® mini kit | 0.9882 | 0.0010 | 0.0033 | 0.0001 | <0.0001 |
| RSC Blood DNA® vs RSC Viral TNA® | >0.9999 | 0.0114 | 0.0155 | <0.0001 | 0.8399 |
| RSC Blood DNA® vs Allprep® DNA/RNA mini kit | >0.9999 | 0.1010 | 0.9988 | 0.0246 | 0.0091 |
| RSC Blood DNA® vs QIAamp® DNA mini kit | 0.2100 | 0.1149 | 0.8099 | 0.3685 | 0.0043 |
| QIAamp® viral RNA mini kit vs RNeasy® mini kit | 0.6184 | 0.6500 | 0.1607 | 0.0018 | 0.0001 |
| QIAamp® viral RNA mini kit vs RSC Viral TNA® | >0.9999 | 0.0055 | 0.0294 | <0.0001 | <0.0001 |
| QIAamp® viral RNA mini kit vs Allprep® DNA/RNA mini kit | 0.9581 | 0.9851 | 0.9197 | 0.9973 | 0.4575 |
| QIAamp® viral RNA mini kit vs QIAamp® DNA mini kit | 0.0512 | 0.0352 | 0.9998 | 0.0080 | 0.0003 |
| RNeasy® mini kit vs RSC Viral TNA® | 0.9973 | 0.0302 | 0.2577 | 0.8465 | 0.0001 |
| RNeasy® mini kit vs Allprep® DNA/RNA mini kit | 0.9992 | 0.0445 | 0.0020 | 0.0018 | 0.0464 |
| RNeasy® mini kit vs QIAamp® DNA mini kit | 0.0643 | 0.0001 | 0.0099 | 0.0001 | 0.0001 |
| RSC Viral TNA® vs Allprep® DNA/RNA mini kit | >0.9999 | 0.0236 | 0.0131 | <0.0001 | 0.0114 |
| RSC Viral TNA® vs QIAamp® DNA mini kit | 0.9407 | 0.0507 | 0.0223 | <0.0001 | 0.0005 |
| Allprep® DNA/RNA mini kit vs QIAamp® DNA mini kit | 0.2013 | 0.0002 | 0.5560 | <0.0001 | 0.0210 |

### Assessment of external conditions

The variability of infection levels in seven sand flies experimentally infected with *L. infantum* is presented in Fig 4. The data presented highlight significant differences in infection intensity among infected individuals, with mean Cq values ranging from 17 to 38, for sand fly DNA extracts obtained using the same assay (EZ1 DSP Virus® Kit). The impact of lyophilization, shipment conditions and presence of sand fly DNA in mean Cq values of EQA samples is depicted in Fig 5A, 5B and 5C, respectively. Lyophilization showed no impact on mean Cq values for low parasite concentrations (10^1^ and 10^2^ for *L. infantum*, 10^1^ for *L. major*). Instead, Cq were significantly greater for higher parasites concentrations (10^3^ for *L. infantum*, 10^2^ and 10^3^ for *L. major*) (Fig 5A). No influences of the storage conditions were noticed, as preservation at -20°C compared to room temperature showed no significant differences in mean Cq values (Fig 5B). Moreover, the presence of DNA from 30 sand flies did not affect *Leishmania* spp. detection at low concentrations. In fact, the efficiency of *L. major* DNA amplification was even better in presence of sand flies (lower Cq values, p-<0.05) (Fig 5C).

**Fig 4.**
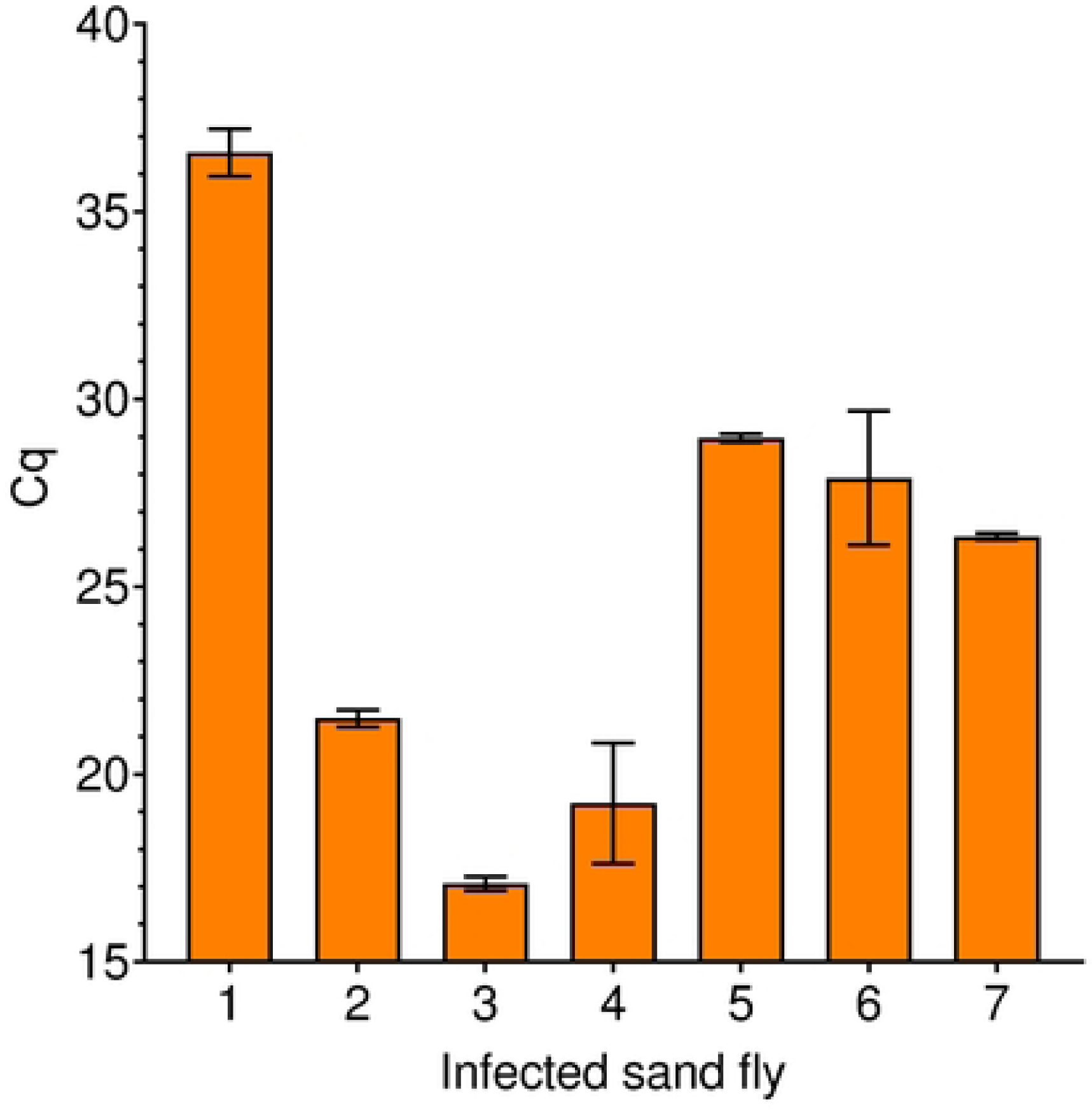
Variability of infection levels in sand flies infected with *Leishmania infantum* under laboratory conditions. Results show mean Cq ± SD of triplicate amplification for each individual sand fly (7 specimens).

**Fig 5.**
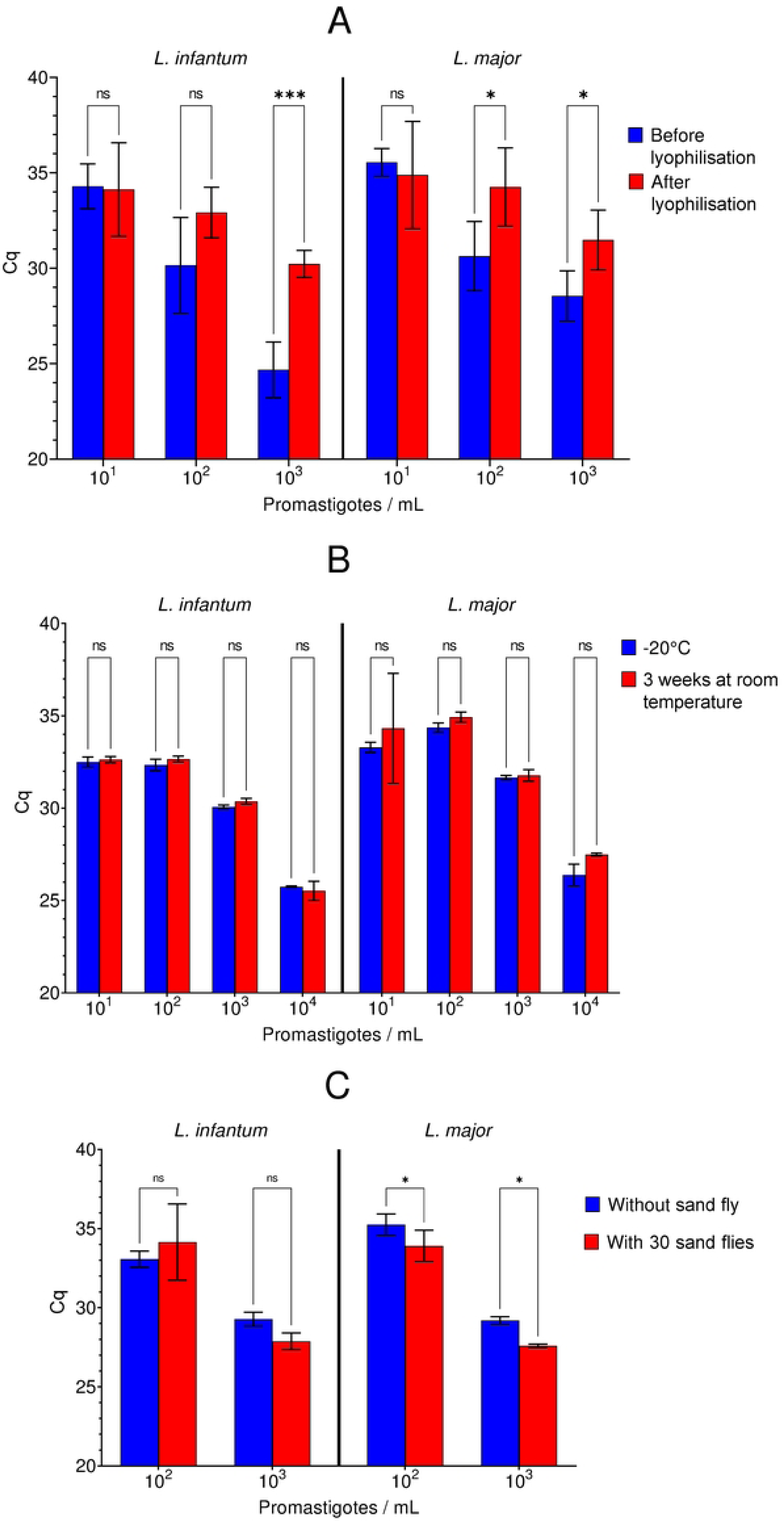
Impact of lyophilization (A), temperature of storage (B), sand fly DNA inhibitors (C) on the efficiency of *Leishmania* DNA amplification. ns: not significant; * p<0.05; ** p<0.01; *** p<0.001.

## Discussion

The implementation of reliable techniques is crucial when they form the core of pathogen surveillance programs, comparing endemicity levels between countries. The CLIMOS project, which aims to combine various parameters (including sand fly density and infection rates), to develop an algorithm for predicting hotspots or the spread of sand fly-borne diseases (SFBDs), must be supported by accurate data. In this context, it was pertinent to implement standard operating procedures to ensure high performance among research centers involved in sand fly and SFBDs surveillance, using the same qPCR amplification method (15) and amplification conditions, to normalize the interpretation of results. However, depending on their own equipment and facilities, the project partners used different extraction methods and qPCR devices. Thus, it was necessary to confirm that centers had comparable performances. With this aim, the Laboratory of Parasitology of Rennes, highly reputed center for evaluation of molecular techniques in the field of human diagnosis (17–19), was in charge of the implementation of an external quality assessment program.

Overall, all centers but one reliably detected all EQA samples corresponding to *L. infantum* and *L. major* concentrations ranging from 10^1^ to 10^4^/mL. The remaining center did not detect *L. infantum* (10^1^/mL) and detected *L. major* (10^1^/mL) only once. Lower sensitivity in qPCR amplification detection could be related to mistakes in sample manipulation, such inaccurate pipetting, or to a comparatively lower performance of the DNA extraction method or of the qPCR master mix used. Unfortunately, it was not possible to retest the 10^1^/mL samples due to lack of DNA. Notwithstanding this, this laboratory was able to amplify the sample containing 10^1^/mL *L. major* and all other samples with higher parasite concentrations. The implications of failing to detect low parasite concentrations are probably low, given that most infected sand flies are likely to contain large parasite numbers, as demonstrated in the present study. The ability of all centers to detect low concentrations of *Leishmania* demonstrates that all partners are duly trained in performing surveillance of *Leishmania*-infected sand flies. Standardization of these methods enables to track changes in infection rates accurately and identify emerging hotspots of transmission. For instance, consistent use of qPCR allows the detection of even low levels of parasite DNA in sand flies, which is critical for early warning and timely intervention in areas where leishmaniasis is spreading. However, the variability in the EQA Cq values provides critical insights into the performance and sensitivity of the diagnostic assay employed. The observed differences in mean Cq values could be attributable to the technical proficiency of the operator, the differences between equipment and positive threshold setting, the DNA extraction kit used, or sample-related factors. The EQA process was evaluated and validated before shipment to ensure reliable comparison of laboratory performances. The variations in DNA yield post-lyophilization could partially explain some discrepancies between centers but would hardly explain the high range of Cq observed for a same concentration. Noteworthy, no significant impact of shipment conditions was observed, thus sample degradation is unlikely to be responsible for the lower sensitivity observed for some laboratories. This finding highlights the need for ongoing training and standardization to ensure uniform diagnostic performance across different laboratories.

All centers were able to detect *Leishmania* DNA in the sample containing a sand fly experimentally infected with *L. infantum*, although differing in quantification cycle (Cq) values. This difference underscores the heterogeneity in host-pathogen interactions at the individual level, even under standardized infection conditions. Indeed, this variability could be the result of fluctuations which are expected and deemed normal within the context of experimental infection, influenced by factors such as the size of the blood meal and the age of the sand fly, among others (20). Despite these variations, the effectiveness of all centers in detecting positive samples was evident, showcasing their proficiency in handling the diagnostic protocols.

After ruling out problems of curve interpretation and Cq threshold variations, extraction methods were evaluated, as a potential source of variability. Results suggest that the choice of the extraction kit may markedly influence the sensitivity of *Leishmania* DNA detection. Taken together, these results may help to understand the discrepancies in Cq values observed between centers. It was observed that the EZ1 DSP Virus® Kit and QIAamp ® DNA mini kit had the best performances for both *Leishmania* species amplification, independently of the parasite concentration. Implementing these kits across laboratories could standardize and improve the consistency and reliability of *Leishmania-*infected sand fly detection. As expected, the use of automated extraction systems, such as the EZ1® robot or Maxwell® device, led to reduced variability, as shown by low error types of triplicate extractions. Despite the superior performance of some DNA extraction kits, it is important to insist that all of them yielded suitable DNA template for effective qPCR detection of *Leishmania*-infected sand flies. However, there remains room for improvement in analyzing low concentrations to ensure high diagnostic validity in *Leishmania* spp. surveillance programs.

As might be expected, the kits developed specifically for DNA extractions (such as EZ1 DSP Virus® Kit, QIAamp® DNA mini kit, Allprep® DNA/RNA mini kit, and RSC Blood® DNA) outperformed those designed for RNA-extraction (such as RNeasy® mini kit and QIAamp® viral RNA mini kit), with avoidance of the DNAase digestion step. These finding implies that field studies aiming at monitoring sand fly-borne infections, *i.e. Leishmania* and phleboviruses, such as CLIMOS, should use a total nucleic acid extraction kit (*e.g.* Allprep® DNA/RNA mini kit) for both pathogens, or use two extraction kits designed for DNA and RNA purification, respectively.

To conclude, all participating centers were proficient in carrying out the diagnostic protocols in the EQA. The detailed comparisons and analyses of different extraction kits for *Leishmania* underscore the importance of selecting the appropriate protocol to ensure high-quality DNA amplification. The benefit of automated extraction, support their adoption across laboratories. While all tested kits are effective, optimizing protocols for low concentration samples remains a key area for improvement to enhance the exhaustive and reliable detection of *Leishmania* in field studies. These results emphasize the importance of standardized protocols and continuous quality assurance to maintain high diagnostic accuracy, which is essential for effective leishmaniasis surveillance in field settings where low concentrations of infection are common.

## Acknowledgment

The authors thank Nazli Ayhan, Rémi Charrel and Laurence Thirion from the UVE laboratory (Unité des Virus Emergents, Marseille, France) for EQA vials lyophilization. They also thank Jovana Sadlova and Petr Volf from Charles University (Department of Parasitology, Prague, Czech Republic) for providing infected sand flies and Jean-Pierre Gangneux (Université de Rennes, Rennes, France) for providing the *Leishmania* strains. And finally, they thank the General Directorate of Public Health/National Parasitology Reference Laboratory (Ankara, Turkey) for local support.

## Author Contributions

F.R-G. and J.P. conceptualized the study, A.D. acquired data on extraction conditions, J.P, A.D. and F.R-G analyzed extraction protocols, F.R-G and J.P. analyzed EQA results, J.P and A.D. wrote the manuscript with the support of F.R-G. J.P., A.D., B.A., U.B., E.B., G.B., J.M.C., M.D.C., T.D.M., O.E.K., E.F., O.D.K., E.K., C.M., M.M., C.M.H., M.N., G.O.K., Y.O., S.O.T., R.P., K.P., C.P., J.R., L.S., G.V., J.W. and K.Y. analyzed EQA. F.R-G edited the manuscript, all authors reviewed and approved the final version of the manuscript.

## Data availability

All resources used in this article are provided in the article and all the analyses are detailed allowing the assessment or verification of the manuscript’s findings.

